# Point-of-care detection of SARS-CoV-2 in nasopharyngeal swab samples using an integrated smartphone-based centrifugal microfluidic platform

**DOI:** 10.1101/2020.11.04.20225888

**Authors:** Ruben R. G. Soares, Ahmad S. Akhtar, Inês F. Pinto, Noa Lapins, Donal Barrett, Gustaf Sandh, Xiushan Yin, Vicent Pelechano, Aman Russom

## Abstract

With its origin estimated around December 2019 in Wuhan, China, the ongoing 2020 SARS-CoV-2 pandemic is a major global health challenge, resulting in more than 45 million infections and 1.2 million deaths. The demand for scalable, rapid and sensitive viral diagnostics is thus particularly pressing at present to help contain the rapid spread of infection and prevent overwhelming the capacity of health systems. While high-income countries have managed to rapidly expand diagnostic capacities, such is not the case in resource-limited settings of low- to medium-income countries.

Aiming at developing cost-effective viral load detection systems for point-of-care COVID-19 diagnostics in resource-limited and resource-rich settings alike, we report the development of an integrated modular centrifugal microfluidic platform to perform loop-mediated isothermal amplification (LAMP) of viral RNA directly from heat-inactivated nasopharyngeal swab samples. The discs were pre-packed with dried n-benzyl-n-methylethanolamine modified agarose beads used as a versatile post-nucleic acid amplification signal enhancement strategy, allowing fluorescence detection via a smartphone camera and simple optics. The platform provided sample-to-answer analysis within 1 hour from sample collection and a detection limit between 100 and 1000 RNA copies in 10 μL reaction volume. Furthermore, direct detection of non-extracted SARS-CoV-2 RNA in nasopharyngeal swab samples from patients with Ct values below 26 (n=25 plus 6 PCR negative samples) was achieved with ∼94% sensitivity and 100% specificity, thus being fit-for-purpose to diagnose patients with a high risk of viral transmission. These results show significant promise towards bringing routine point-of-care COVID-19 diagnostics closer to resource-limited settings.

## 1. Introduction

Simple, rapid, and sensitive analytical methods for viral detection at the point-of-care have been increasingly in demand in recent decades. Such devices are especially relevant in resource-limited settings (RLS) in Africa, Asia and South America, where the prevalence of ubiquitous viral pathogens such as Human Immunodeficiency Virus (HIV) and other tropical viruses including Zika, Ebola, Crimean-Congo Haemorrhagic Fever (CCHF), Chikungunya and Dengue claim tens of thousands of human lives per year mostly due to the lack of effective diagnostics and subsequent disease containment and therapy ^1-3^. However, few occasions in modern history have been more pressing for developments in this field than the present SARS-CoV-2 coronavirus pandemic ^4^ which from December 2019 to October 2020 has infected more than 45 million people in over 214 countries and territories, resulting in more than 1.2 million deaths. Both these figures are still growing rapidly.

The first cases of SARS-CoV-2 infection were reported in December 2019 in Wuhan with a probable zoonotic origin considering its genome sequence is closely related to bat coronaviruses ^5^. Although other human coronaviruses associated with epidemic outbreaks, i.e. SARS-CoV and MERS-CoV, had a higher mortality rate, the much higher infectivity and long asymptomatic incubation period associated with SARS-CoV-2 infections, resulting in the pathologic condition designated as Corona Virus Disease-19 (COVID-19), allowed the rapid spreading of the virus around the globe ^6^. In extreme cases, a detectable viral load in nasopharyngeal swabs was observed prior to symptom onset ^7^ and as long as 20 days after the first symptoms ^6^. Furthermore, considering the significant differences in symptoms and their severity, it is estimated that as many as 90% of all infections were undocumented early in the pandemic before travel restrictions were imposed ^8^.

Aiming at controlling the spread of the virus, millions of viral load tests have been performed in centralized labs supported by sample collection in the field to screen symptomatic and asymptomatic infections. The sampling methods of choice are nasal or throat swabs stored in viral collection medium prior to RT-PCR ^9^, with primers/probes targeting the ORF1ab, RNA dependent RNA polymerase (RdRp), envelope (E), spike protein (S) or nucleocapsid (N) genes of the SARS-CoV-2 single stranded RNA genome ^10-12^. Viral load values during the course of the infection typically range between ∼10^3^ to ∼10^11^ copies/mL ^6,7,9,13^ and > 10^6^ copies/mL are common early upon the onset of symptoms ^7^, with a peak in the order of 10^9^ copies per throat swab on day 4 after onset of symptoms ^14^. However, in the context of viral viability and risk of transmission, it has been observed that the probability of isolating infectious SARS-CoV-2 is less than 5% when the viral load in the respiratory tract is below ∼4.3×10^6^ RNA copies/mL ^15^. Thus, it has been recently advocated that more frequent and cost effective testing with lower sensitivity, instead of lengthy and sparse PCR testing in centralized labs, can provide a more efficient strategy for containment by (1) rapidly detecting the viral load peak early upon infection and (2) avoiding unnecessary isolation during the long viral load decrease period detectable only with high analytical sensitivity and with minimal to zero risk of cross-human infection ^16^. In this context, it is expected that nucleic acid amplification tests can still provide higher sensitivity and potentially specificity than antigen rapid tests ^17^, thus being the ideal solution if costs and equipment complexity can be kept low.

Within the landscape of the ongoing pandemic, it has become clear that testing is paramount to reduce the spread of infection and ease the impact on health systems combined with a smart and data-driven management of social distancing policies. However, while developed countries can economically and logistically support an exponential increase in lab-based testing and social distancing measures, such a course of action is hardly feasible in RLS ^18^. Thus, to support routine SARS-CoV-2 diagnostics on a global scale but particularly in RLS, a cost-effective, rapid, sensitive, portable and simple to use device to detect RNA directly from biological specimens would have a significant impact.

Concerning the development of portable analytical devices, the use of RT-PCR is suboptimal considering the intrinsic technical complexity of performing several precise heating-cooling cycles up to >94 °C ^19^. Thus, in the context of pathogen detection, several groups have been actively exploring the use of loop-mediated isothermal amplification (LAMP) as a potential alternative, requiring only a constant and relatively lower temperature of ∼65 °C ^19^. Concerning the miniaturization of LAMP towards integrated analytical platforms, its combination with centrifugal microfluidics ^20-24^ and/or smartphone-based signal readout ^25-32^ has been showing significant promise to achieve a true sample-to-answer operation. A few remarkable examples of LAMP-based miniaturized modules using either of these approaches are: (1) capture of LAMP amplification products with anti-DIG antibodies followed by generation of a TMB precipitate measured using a standard light source and a smartphone camera ^25^; (2) digital microfluidic platform with temperature monitoring/control provided by a thermal imaging camera and SYBR Green I derived fluorescence transduction by naked eye or smartphone camera ^27^; (3) Microfluidic cartridge combining immune-capture, lysis and LAMP to detect viable bacteria using a reader platform comprising two light sources for fluorometric and/or turbidimetric analysis resorting to a smartphone camera ^30^; (4) a hermetic container providing power-free chemical-based heating for LAMP amplification followed by detection using a smartphone flashlight and camera for fluorometric detection ^32^; (5) Centrifugal platform combining silica-based DNA extraction and integrated LFA strips to multiplex the detection of multiple LAMP products using anti-DIG antibodies and colorimetric detection ^32^; (6) Centrifugal platform with automated bead-beating lysis followed by direct RT-LAMP by continuous measurement of fluorescence with UVC illumination and a standard camera ^22^; and (7) Centrifugal platform incorporating non-contact heating of the disc and colorimetric detection of LAMP products using a white LED for illumination and filtered photodiodes for signal acquisition ^24^.

Here, LAMP, centrifugal microfluidics, smartphone-based detection and recent developments in RT-LAMP applied to the detection of SARS-CoV-2 RNA ^33-39^ are combined to develop a novel cost-effective and fully integrated platform for COVID-19 diagnostics directly from heat-inactivated nasopharyngeal samples. The direct detection from heat-inactivated samples was achieved using (1) a one-pot combination of reverse transcriptase and polymerase enzymes for robust isothermal amplification and (2) a novel agarose bead-based signal enhancement strategy for improved fluorometric detection, thus avoiding the impact of collection media on weakly-buffered pH responsive colorimetric amplification mixtures ^39^.

## 2. Results and Discussion

### 2.1. Characterization of RT-LAMP sensitivity and development of a bead-based strategy for fluorescence signal enhancement

Considerable advances in LAMP-based detection of SARS-CoV-2 RNA have been achieved since the beginning of the pandemic, with several primer sets and potential detection strategies being reported in the past 6 months. In this context, particular attention has been given to pH-based colorimetric signal transduction considering the simple visual interpretation of the results ^39^. However, mildly buffered systems are highly prone to interference from different biological samples and collection media, typically requiring a previous silica-based extraction and elution with DI-water for improved robustness ^39^. Fluorescence detection using DNA intercalator dyes is an equally simple and more interference-forgiving approach but requires the tackling of two key limitations, namely (1) the relatively more complex signal transduction, particularly in miniaturized systems concerning low fluorescence signal intensities and filtering requirements ^40^ and (2) the high numbers (typically 6 different sequences) and design complexity of LAMP primers results very often in a variable degree of intra- (hairpins) and cross-primer hybridization (primer dimers), which result in significant non-specific fluorescence signal ^41^. This signal is particularly relevant at room temperature due to the lower hybridization stringency, hindering a simple, non-real-time, post-LAMP measurement. Here, the developed device incorporates an agarose bead-based strategy combined with centrifugal microfluidics to fundamentally tackle both these limitations by significantly enhancing the negative-to-positive signal ratio. Using a set of primers that we have previously developed ^33^ and resorting to a combination of SSIV and Bst 3.0 for LAMP and SYBR Green I for fluorescence generation, the obtained RT-LAMP results are shown in **Figure 1-A**. Performing a dilution series of an RNA fragment with a 226 bp sequence matching the ORF1ab of SARS-CoV-2 in water, 100 copies per 10 μL reaction could be detected within 30 min of amplification time, in agreement with the typical LoD requirements of FDA-approved COVID-19 diagnostic technologies ^4^. Strikingly, while the increase in fluorescence signal magnitude from negative to positive measured at 65 °C was about 5-fold, the difference was reduced to ∼1.5-fold at room temperature (∼25 °C) due to an increase in signal in the non-amplified samples. This background, arising from the increased hybridization of primer dimers/hairpins at lower temperatures, was addressed using the novel bead-based sample processing strategy described in **Figures 1-B** and **1-C**. This strategy dramatically improved the signal-to-noise ratio at room temperature, thus facilitating signal acquisition when portability and cost reduction are a priority. Multimodal ligands as is the case of N-benzyl-N-methylethanolamine (NBNM) combining anion exchange and other interactions, i.e. hydrophobic and hydrogen bonding, are typically used in chromatography to capture host cell DNA as an impurity ^42^ or plasmid DNA as target product ^43^ directly from complex matrices. Here used for sample processing, this ligand, when modified on agarose beads with an average porosity of ∼ 100 nm, was found to have a remarkable selectivity for short oligonucleotides with lengths of 25-100 bp and a cutoff region between 200-400 bp (**Figure 1-B**). According to the results in **Figure 1-B**, 1.3 μL of beads could deplete virtually all the primers in 10 μL of LAMP mixture (Negative Input vs Negative Output), while the titers of LAMP products amplified from 10^6^ copies of template RNA remained constant above 400 bp (Positive Input vs Positive Output). These observations were confirmed using fluorescence microscopy in the presence of SYBR Green I as dsDNA selective intercalator. According to the results in **Figure 1-C**, the primers in a LAMP mixture without RNA template (Negative) result in a very high fluorescence measured at bead level, which is ∼4.6-fold higher compared to a positive LAMP mixture (10^3^ copies of RNA template). This observation is in agreement with the electropherograms in **Figure 1-B** which highlight the complete depletion of all primers (< 50 bp) in the presence of template (Negative Input vs Positive Input). The fluorescence microscopy measurements also highlight an increase of ∼28-fold in signal magnitude between negative and positive, comparing the measurement performed directly on solution before bead processing (upstream of the beads) relative to the measurement performed directly on the beads. Such an increase in signal magnitude shows promise to implement a robust signal acquisition strategy with minimal instrumentation complexity. The versatility of the bead-based enrichment as a general post nucleic acid amplification signal amplification strategy was further validated for up-concentrating of SARS-CoV-2 PCR amplicons (100 bp long) detected with a molecular beacon (**Figure S1**). In agreement with the characterization in Figure 1-B, the short PCR amplicons were effectively captured within the pores of the NBNM beads, contrarily to the longer LAMP products, thus generating a linear and directly proportional correlation between bead fluorescence and RNA copies with Ct ranging from 25 to 36 (R^2^ =0.99) (**Figure S1**). Schematics illustrating the bead capture behavior in the presence of LAMP or PCR positive and negative samples are shown as supplementary information (**Figure S2**).

**Figure 1.**
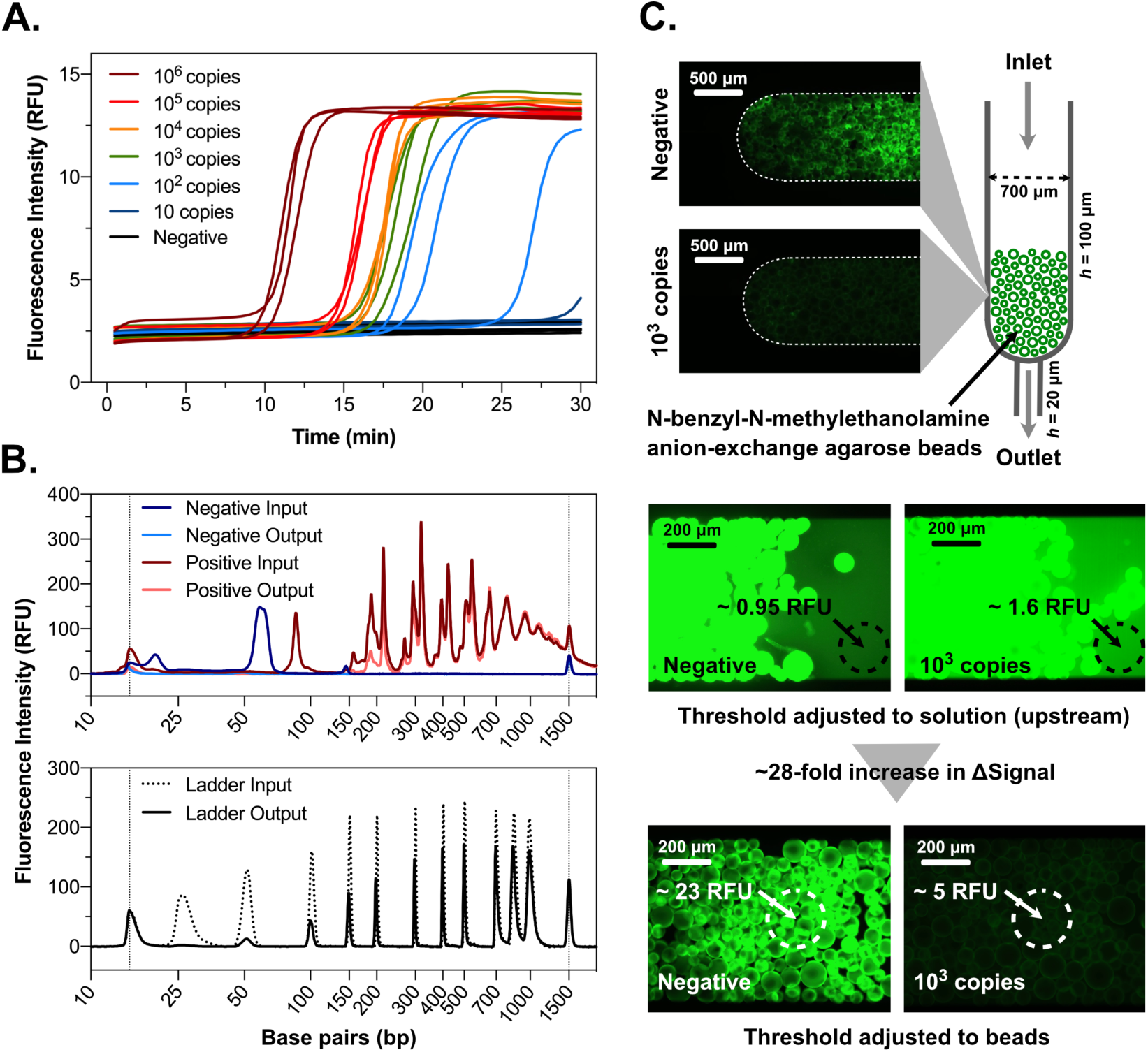
Characterization of LAMP assay and bead-based signal enhancement. **A**- Dilution series of SARS-CoV-2 RNA fragment spiked in water. The RNA copies were added to a 10 μL reaction mix and measured in triplicate. **B (top)**- Capillary electropherograms of the LAMP master mixes incubated at 65 °C for 30 min in the presence (positive) or absence (negative) of 10^6^ SARS-CoV-2 RNA fragment copies before (input) and after (output) processing using NBNM beads packed on a disc. **B (bottom)**- Capillary electropherogram of a DNA ladder (25-1,000 bp) before (input) and after (output) processing using NBNM beads packed on a disc. Peaks below and above 25 and 1000 bp, respectively, correspond to the boundaries of the electropherogram. **C**- Fluorescence microscopy of NBNM beads packed on a PDMS microcolumn after flowing a LAMP mixture pre-incubated at 65 °C for 30 min in the presence (positive) or absence (negative) of 10^3^ SARS-CoV-2 RNA fragment copies. The signals were measured as average grayscale intensities on the highlighted regions. ΔSignal refers to the difference in signal magnitude between the positive and negative sample measured on the solution upstream of the beads (0.65 RFU) and directly on the beads (18 RFU).

### 2.2. Development of an integrated platform for point-of-care COVID-19 diagnostics using a smartphone camera for fluorometric signal transduction

Aiming at bringing cost-effective, rapid and sensitive COVID-19 diagnostics to RLS, the main goal of this work was to develop a robust but minimally complex and portable platform for SARS-CoV-2 RNA detection directly from heat inactivated biological samples. Taking advantage of the signal enhancement strategy reported in section 2.1 the concept was based on designing a PMMA disc with 20 independent channels (**Figure 2-A**) each comprising (1) a region to perform LAMP of 10 μL sample (LAMP section), (2) a section containing packed NBNM beads at the interface between 400 and 50 μm deep channels and (3) an outlet chamber (OC) connected to the previous two components via the narrow 50 μm deep section. To process the sample added to the disc, the sequence of steps schematized in **Figure 2-B** was followed. The heat-inactivated swab sample containing template RNA was mixed with the LAMP reagents and a total of 10 μL of the mixture was added to the channel, the access holes were sealed and the whole disc incubated at 65 °C. After amplification, the disc was subjected to four cycles of ramping the rotation linearly from 0 to 6,000 rpm and vice-versa (1 cycle meaning 0-6,000-0 rpm). During the ramp-up, the liquid is forced through the beads due to the centrifugal force and held back by an increase in pressure inside the sealed OC. During ramp-down, the decrease in centrifugal force allows the OC to decompress pushing the liquid backwards, while the denser agarose beads are still held in place by the centrifugal force. This back and forth motion with 4 cycles allows a complete capture of the primers in solution according to the previous results in **Figure 1-B**.

**Figure 2.**
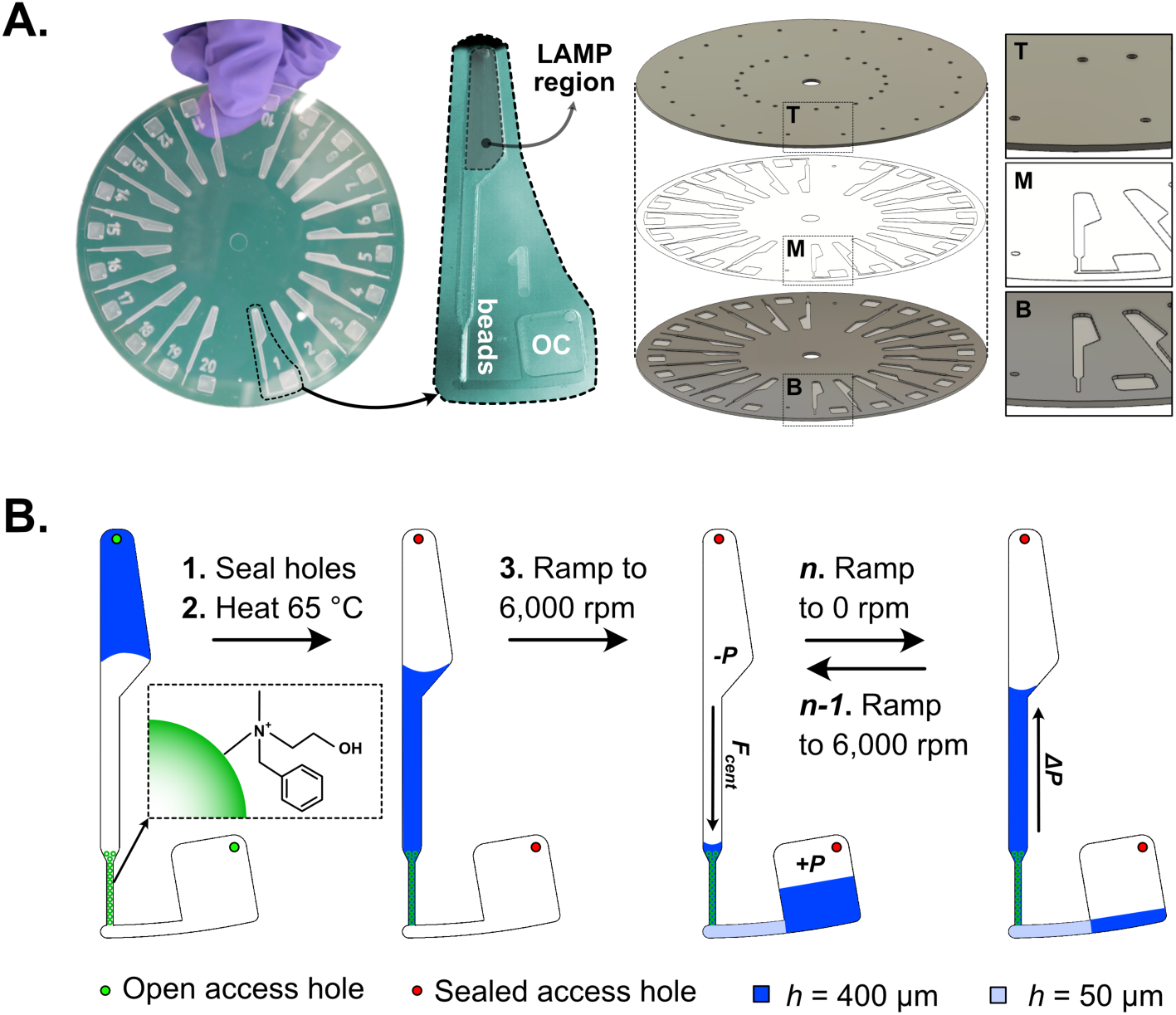
Schematics and working principle of the on-disc LAMP fluorescence signal readout with bead-based signal enhancement. **A**- PMMA disc comprising 20 parallel channels packed with dried NBNM agarose beads. The disc is fabricated with 3 layers, a bottom (B) 1 mm PMMA layer with embedded 400 μm deep channels, a middle (M) layer comprising patterned double-sided PSA defining a 50 μm deep sieving channel preventing the flow of beads into the outlet chamber (OC), and a top (T) 1 mm PMMA layer with inlet and outlet access holes. **B**- Sequential operation of the disc after adding 10 μL of sample. The sequence comprises the (1) sealing of the inlet and outlet holes, (2) LAMP by heating the disc at 65 °C for 30 minutes, (3) ramping up the rotation speed to 6,000 rpm to force the solution through the packed beads, followed by a ramp down to 0 rpm, resulting in a backflow of the liquid due to the pressure difference between the OC (positive pressure) and the LAMP region (negative pressure). The final two steps can be repeated multiple times to ensure complete capture of the target molecules in solution.

The portable platform developed to combine the centrifugal and heating modules required to perform the LAMP followed by the bead-based signal enhancement is shown in **Figure 3**. The top centrifugal module shown in **Figure 3-A** uses a DC motor controlled by a microcontroller board and a hall-effect sensor to complete the rotation protocol. After the rotation protocol, the measurement is performed through a PMMA lens in the cover lid covered on the backside with a 100 μm polyimide film which is aligned with the smartphone camera with a custom-made adapter. The polyimide film serves as emission absorption filter to block the excitation light from the 450 nm 80 mW laser embedded on the platform ^44^. The laser diode is aligned at an angle of ∼ 17.5° relative to the disc (**Figure S5**) to take advantage of total internal reflection of the PMMA-air interface between the disc and the lens to minimize residual leakage of blue light into the camera sensor.

**Figure 3.**
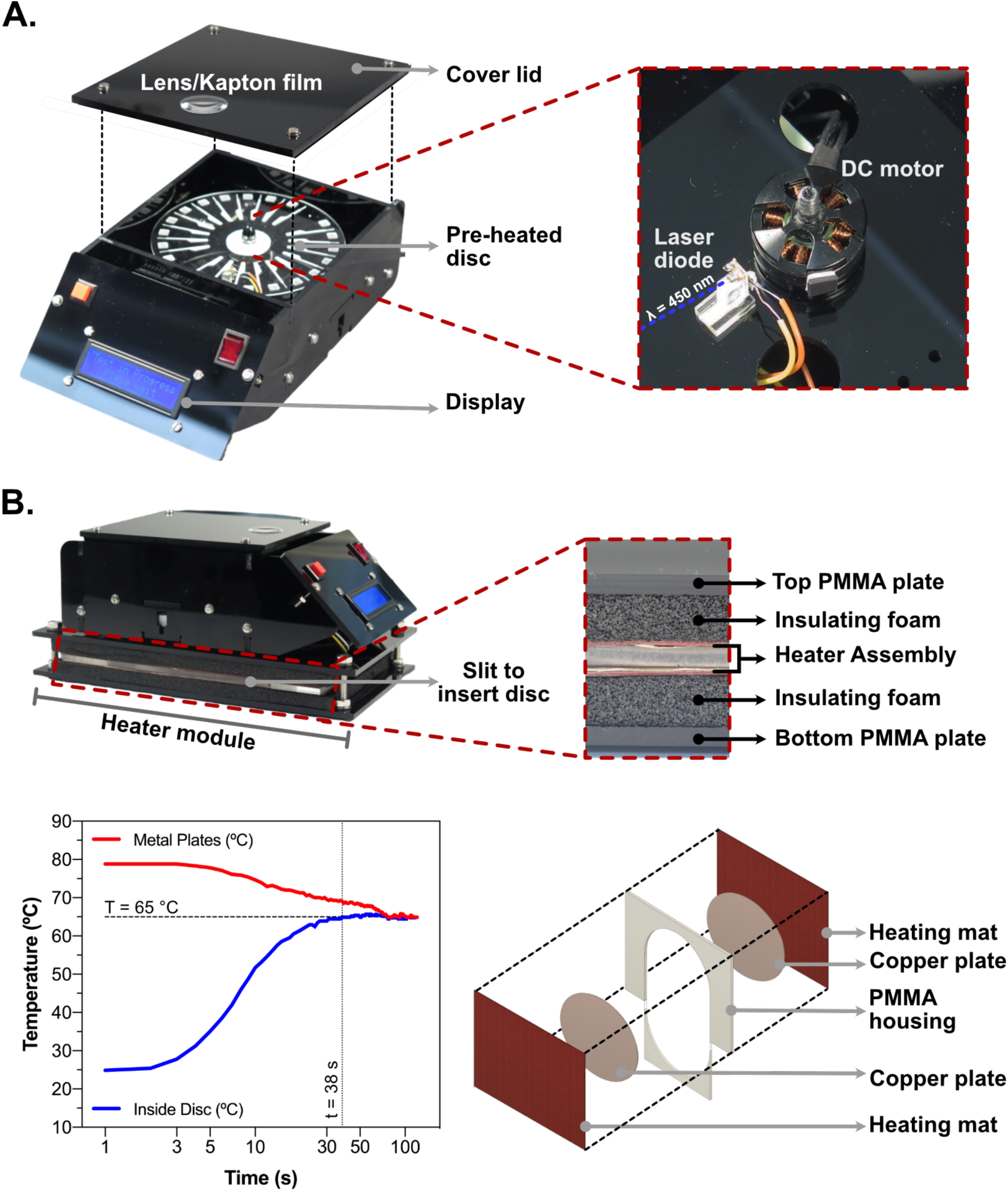
Platform combining centrifugal and heating modules for LAMP-based diagnostics coupled with fluorometric readout using a standard smartphone camera. **A**- Photographs of the key components of the centrifugal module. The bottom side of the PMMA lens is covered with a Kapton (polyimide) film working as emission filter. **B**- Photographs and schematics of the key components of the heating module. The disc is enclosed in a PMMA housing and in between two copper plates to maximize the rate of heat transfer and maintain a constant temperature. The starting temperature of the copper plates (continuously measured between the plates and the heating mats) is set above 65 °C to strike a balance between maximum heating rate inside the disc channels (38 s to reach 65 °C) while preventing overshooting.

The heating module assembled below the centrifugal platform is shown in **Figure 3-B**. This module comprises a PMMA housing with a removable front piece to insert the disc. The disc enclosed by the housing is sandwiched in contact with two copper plates which are actively heated by two resistive silicone mats. The temperature control is achieved with a feedback loop measuring the temperature between the copper plates and the heating mats on both sides of the stack. The PMMA housing and the insulating foam on each side of the heating mats serves to minimize convective and conductive heat dissipation. To minimize non-specific amplification of hybridized primer dimer and primer-template pairs at lower temperatures, rapid heating is achieved by initially setting the temperature of the copper plates at 79 °C according to the plot in **Figure 3-B**. The plates are passively cooled down by the disc and active heating is only actuated once the temperature of the plates decreases below 65 °C.

### 2.3. Characterization of the integrated platform and benchmarking against RT-LAMP

The developed platform was subsequently characterized by testing a dilution series of ORF1ab RNA fragment spiked in DI-water (**Figure 4**). A dilution series using the complete platform was tested and the results in **Figure 4-A** show a limit of detection between 10^2^-10^3^ copies/reaction (0% positive at 10^2^ copies and 100% positive for 10^3^ copies performing 4 independent measurements for each concentration). The analysis of the signal in each channel was performed by measuring the grayscale intensity profile of the green channel (average of 30 pixels) 100 pixels along the interface of the beads and the solution. Centering the 30×100 pixels line profile at the bead-solution interface (negative pixel values −50 to 0 in solution and positive values 0 to 50 on the beads), the response was described as a 10-pixel moving average relative to the intensity of the solution. A negative/positive decision threshold was defined according to 3.29 times the standard deviation of the relative signal obtained at the end of the line profile (pixel 50) for 5 independent negative controls. The obtained sensitivity for the on-disc LAMP was lower than that obtained in the RT-LAMP tests (**Figure 1-A**), where 100% of the samples with 10^2^/copies per reaction showed a positive signal. The relatively lower sensitivity is hypothesized to arise from non-specific adsorption of enzymes and/or template to the PMMA channel, having a significantly higher surface area in contact with the 10 μL of solution compared to a standard reaction tube. Efforts to improve channel passivation strategies or application of alternative materials are envisioned to maximize performance. **Figure 4-B** highlights the enhanced signal (Negative vs 10^3^ copies of target RNA fragment) provided by the beads. While for the previous fluorescence microscopy analysis discussed in section 2.1 the focus was placed on the beads and LAMP solution before processing, the photographs shown here highlight also the complete removal of non-specific fluorescence signal in solution for the negative control. These observations support the combination of both types of measurements, i.e. directly on the beads and on the solution, to allow the maximization of the signal-to-noise ratio. The bead-based primer depletion was also validated as a simultaneous means of inactivating the reaction post-LAMP, thus avoiding higher temperatures and longer assay times for enzyme inactivation (**Figure 4-C)**. It was observed that when the primer depletion cycles were performed before amplification (65 °C for 25 min), no LAMP products were obtained with initial RNA template titers as high as 10^9^ copies/reaction. Overall these results confirm the triple functionality of the agarose beads for (1) signal intensity enhancement, (2) sample preparation to remove non-specific background and (3) simple reaction inactivation at room temperature.

**Figure 4.**
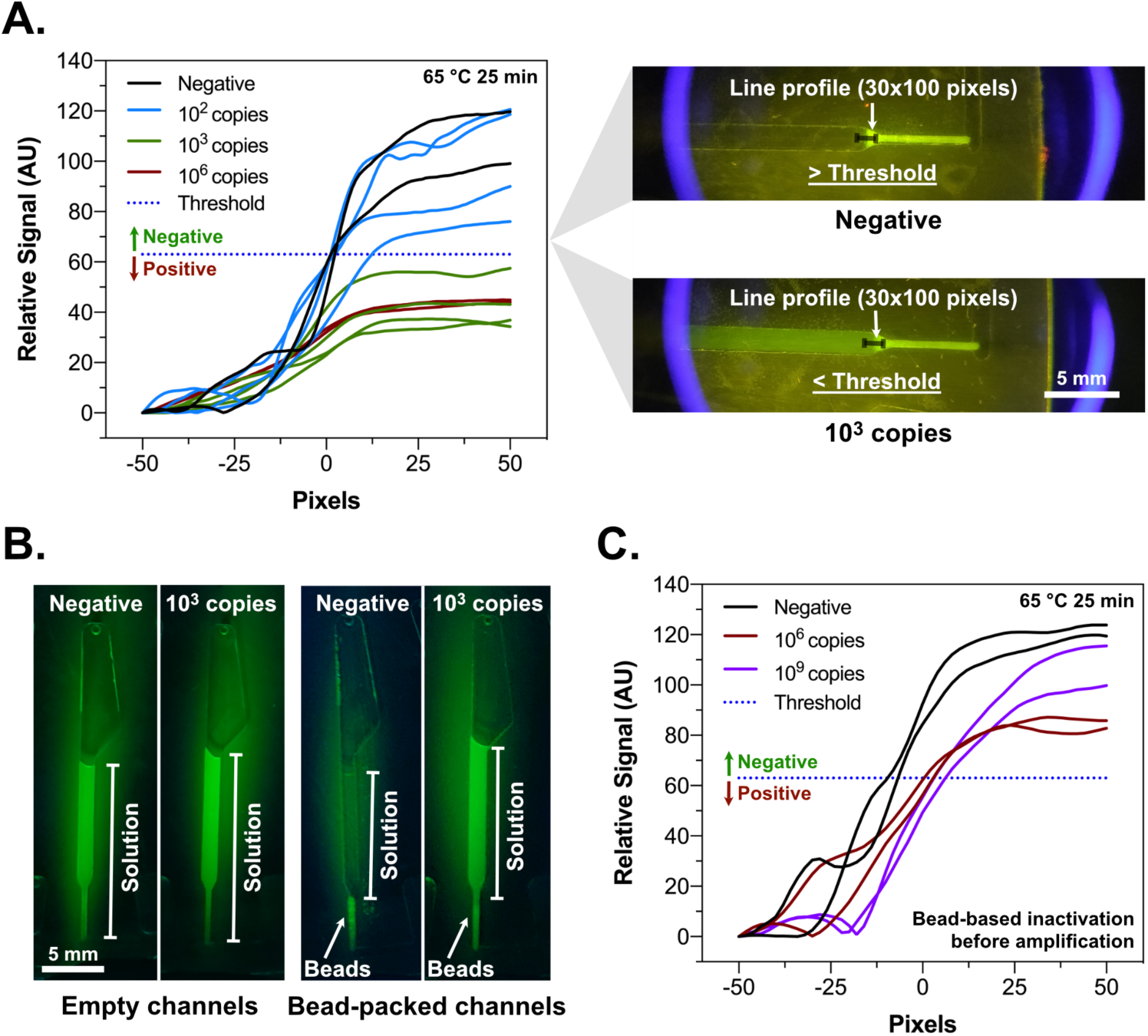
Characterization of on-disc signal generation after on-disc LAMP. **A**- Relative grayscale intensity (green channel) measured at the interface between the solution and beads (50 pixels each side) performing the LAMP in the presence of increasing copy numbers of SARS-CoV-2 RNA fragment. The threshold value was calculated as the average difference between signal magnitude of the beads and solution minus 3.29 times the standard deviation of 5 independent negative measurements. Differences in signal magnitude at pixel 50 above and below the threshold are considered negative and positive, respectively. **B**- Enhancement of signal-to-noise ratio in solution provided by the NBNM beads. All images (green channel only) were acquired using a smartphone camera combined with illumination provided by the 450 nm laser diode and polyimide film as emission filter. **C**- Bead-based LAMP inactivation. The LAMP was performed on the disc after first flowing the mix through the NBNM beads according with the same rotation protocol used for the measurements.

### 2.4. Direct detection of SARS-CoV-2 in heat-inactivated nasopharyngeal swab samples

The developed platform was further validated for the detection of SARS-CoV-2 viral RNA directly from heat inactivated (95 °C, 15 min) nasopharyngeal swab samples collected in two different types of storage media, namely Sigma Virocult^®^ and Sigma Transwab^®^. A total of 31 clinical samples summarized in **Table 1** were tested and pre-analyzed by RT-PCR targeting the E and N genes. All samples were first tested in duplicate using RT-LAMP and the results are compiled in **Figure 5-A**. Overall, for the tested samples, 100% sensitivity and specificity were achieved after 25 min incubation time, with a positive signal obtained within 20 min for Ct values as high as 27.0 and no amplification was obtained for the negative samples. Furthermore, a good correlation was obtained between the Ct values of RT-PCR and the time to positivity (TtP) of RT-LAMP (Pearson correlation coefficient of 0.89). To maximize sensitivity when testing the samples using the on-chip LAMP, an incubation time of 30 min was necessary as demonstrated in **Figure 5-B**. Focusing on a set of 2 samples in each Ct range, i.e. Ct < 15 (V), 15 < Ct < 20 (H), 20 < Ct < 25 (M), Ct > 25 (L) and PCR negative (N), an increase in amplification time from 25 to 30 min shifted the sensitivity by at least 5 Ct values from the H to the L range. Longer incubation times were not tested to prevent false positive signals. The output of the platform for the analysis of all samples listed in Table 1 is compiled in **Figure 5-C**. The plotted signals are defined as the signal magnitude measured 50 pixels into the region containing the packed beads relative to the signal measured 50 pixels towards the solution in accordance with the extremes of the profile illustrated in **Figure 5-B**. Among the tested samples, 100% sensitivity and specificity were obtained for samples with Ct values < 25 and ∼94% sensitivity/100% specificity including samples in the high Ct range (L1-L3). Overall, the developed platform allowed the detection of SARS-CoV-2 RNA with Ct values as high as 26.1 directly for heat-inactivated nasopharyngeal samples within 40 min from sample inactivation to result.

**Table 1.** Nasopharyngeal swab samples analysed using Cepheid GeneXpert targeting E and N genes. “Average Ct” refers to the average of both E and N gene Ct values. “Medium” refers to the collection tube used to store the nasopharyngeal swab samples prior to analysis. Virocult tubes contained liquid Virocult medium and Transwab tubes contained liquid Amies medium.

| Sample ID | Ct E | Ct N2 | Average Ct | Medium |
| --- | --- | --- | --- | --- |
| V1 | 11 | 13.5 | 12.3 | Virocult |
| V2 | 12.2 | 14.5 | 13.4 | Virocult |
| V3 | 13.5 | 15 | 14.3 | Transwab |
| V4 | 13.3 | 15.8 | 14.6 | Transwab |
| V5 | 13.7 | 15.4 | 14.6 | Transwab |
| H1 | 14.3 | 16.3 | 15.3 | Virocult |
| H2 | 15.6 | 17.8 | 16.7 | Virocult |
| H3 | 16.9 | 18..8 | 16.9 | Transwab |
| H4 | 17.1 | 18.9 | 18.0 | Transwab |
| H5 | 17.6 | 19.8 | 18.7 | Virocult |
| M1 | 19.2 | 21.2 | 20.2 | Virocult |
| M2 | 19.4 | 21.6 | 20.5 | Virocult |
| M3 | 20.1 | 22.3 | 21.2 | Transwab |
| M4 | 20.1 | 22.5 | 21.3 | Transwab |
| M5 | 20.6 | 22.7 | 21.7 | Virocult |
| M6 | 20.8 | 23.2 | 22.0 | Virocult |
| M7 | 21.3 | 24.2 | 22.8 | Virocult |
| M8 | 21.6 | 24.2 | 22.9 | Transwab |
| M9 | 22.2 | 24.2 | 23.2 | Virocult |
| M10 | 22.1 | 24.7 | 23.4 | Transwab |
| M11 | 22.5 | 24.8 | 23.7 | Transwab |
| M12 | 23.4 | 25.3 | 24.4 | Transwab |
| L1 | 25.1 | 27.1 | 26.1 | Transwab |
| L2 | 25.1 | 27.4 | 26.3 | Transwab |
| L3 | 25.7 | 28.2 | 27.0 | Transwab |
| N1 | NA | NA | NA | Virocult |
| N2 | NA | NA | NA | Virocult |
| N3 | NA | NA | NA | Virocult |
| N4 | NA | NA | NA | Virocult |
| N5 | NA | NA | NA | Transwab |
| N6 | NA | NA | NA | Transwab |

**Figure 5.**
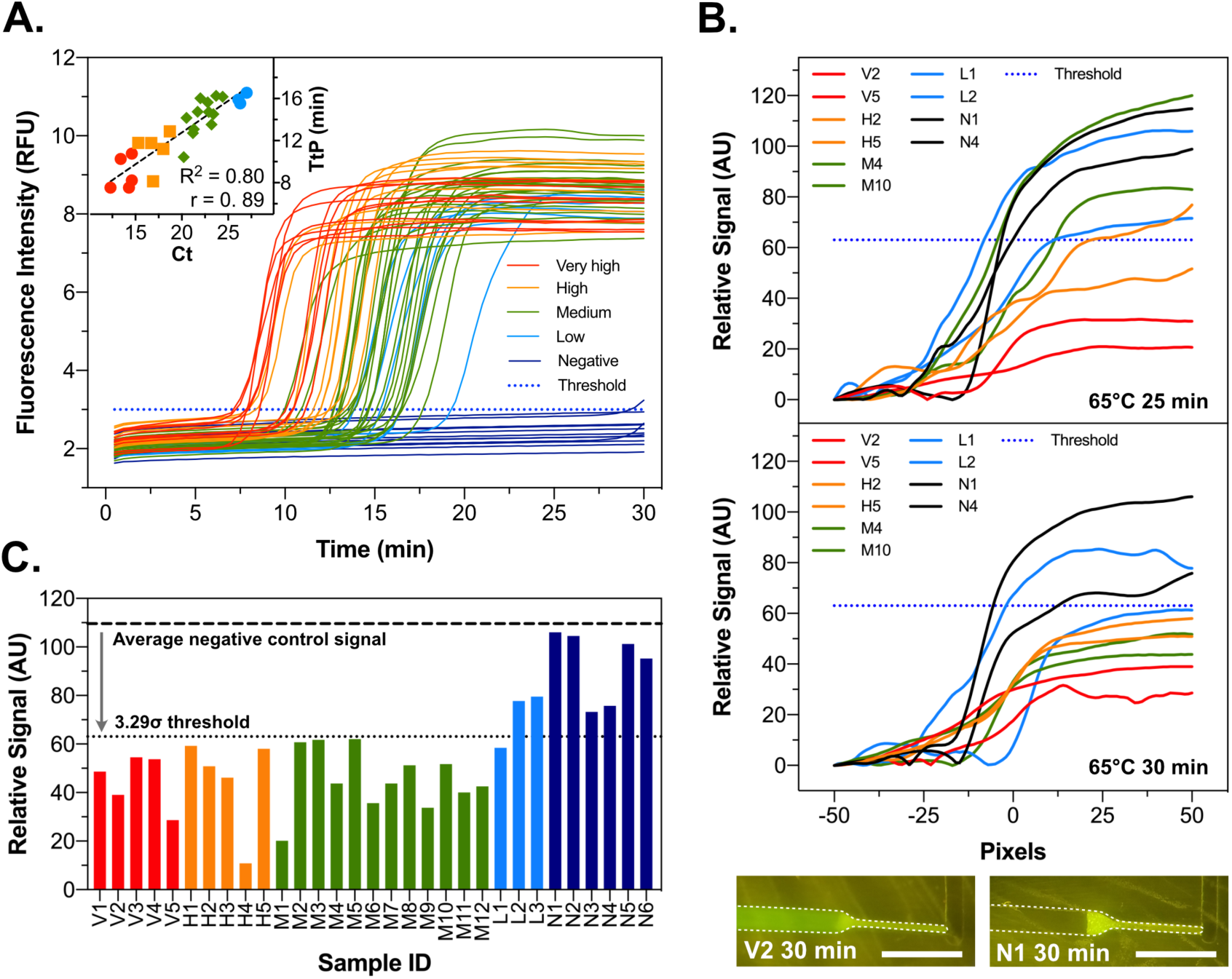
Detection of SARS-CoV-2 in heat inactivated nasopharyngeal swab samples. The samples in Table 1 were grouped in 5 categories, namely (1) very high – average Ct <15 (red), (2) high – Ct between 20 and 15 (yellow), (3) medium – Ct between 20 and 25 (green), (4) low – Ct > 25 (blue) and (5) negative (dark blue). Color coding applies to A, B and C. **A**- RT-LAMP analysis of clinical samples using a benchtop real-time thermocycler (micPCR). All samples were measured in duplicate. The inset plot shows the correlation between average PCR Ct value of each sample and time to positivity (TtP) measured for RT-LAMP. TtP was determined as the required incubation time at 65°C to increase the fluorescence intensity above the threshold. The threshold was fixed as the highest background fluorescence (before amplification) measured among all tested samples. **B**- Analysis a set of 2 samples in each Ct range using the integrated on-chip platform with an incubation time of 25 or 30 min at 65°C. Scale bars: 5 mm. **C-** Relative signal measured at pixel 50 for each nasopharyngeal swab sample. Relative values below the threshold are considered positive for SARS-CoV-2 RNA. The threshold and relative signal values in B and C were determined as previously described in section 2.3.

## 3. Conclusions

We introduce a novel portable bead-based centrifugal microfluidic platform and demonstrate LAMP based viral RNA detection directly from heat-inactivated nasopharyngeal swab samples. The platform achieves two major breakthroughs in the scope of point-of-care viral diagnostics. Firstly, a versatile agarose bead-based strategy was developed to significantly improve signal transduction after LAMP by removing the intrinsic background of primer dimer interactions when using intercalating fluorescent dyes. Secondly, the developed centrifugal and heating modules serve as a complete package for sample-to-answer analysis directly from heat-inactivated nasopharyngeal samples in less than 1 hour of total processing time, combined with simple smartphone-based signal acquisition. The limit of detection of ∼26 Ct (approximately 1×10^4^-1×10^5^ copies/mL ^45^) and inexpensive analysis are a suitable combination for frequent screening to allow detection of the spike in viral load early upon infection and minimize the risk of transmission ^16^. In the context of the current SARS-CoV-2 pandemic, the full centrifugal platform is adaptable to any smartphone, costs less than 200 USD and is compatible with cost-effective and scalable LAMP reagents under investigation ^46^. These features can potentially pave the way to bring routine and scalable diagnostics to RLS, as well as expanding the current diagnostic capacities in high-income countries by bringing viral RNA detection directly to the field. Both these avenues are paramount to improve the containment of viral spread and simultaneously minimize the need for extreme lockdown policies.

## 4. Methods

### 4.1. Fabrication of PDMS microchannels and discs

The PDMS microchannels used for fluorescence microscopy characterization were fabricated as described in detail elsewhere ^47^. Briefly, microchannels comprising a 700×100 μm^2^ cross section converging into a 200×20 μm^2^ cross-section aimed at trapping agarose beads with an average diameter of 90 μm, were fabricated using standard SU-8 mold replication techniques. 18-gauge inlet and outlet access holes were punched using a blunt syringe and the channels were sealed against Corning glass slides after an oxygen plasma treatment (30 sec, Femto Science CUTE, 100 W, 80 Pa O_2_).

The discs were designed using Autodesk® Fusion 360 (Education License) and cut into 1 mm thick PMMA sheets using computer-numerical-control milling machine (Roland Modela MDX-40A). The disc layers were bonded together using clear medical grade pressure-sensitive adhesive (ARcare® 92712). The microchannels on pressure-sensitive adhesives were cut using a cutter plotter (Graphtec CE6000-40). After aligning the PMMA layers with the pressure-sensitive adhesive layer, the disc was placed in a manual press machine overnight to ensure uniform bonding. Detailed dimensions of each microchannel are shown in **Figure S3**.

### 4.2. Assembly of the heating module

The heating module consisted of two silicone heater mats (30 W, 150 × 200 mm, 12 V DC; RS PRO Article # 731-366) which were used to heat up 1 mm thick copper plates (120 mm diameter.). The heater mats are self-adhesive and the copper plates were attached to the mats. A 4 mm thick PMMA sheet served as a chamber for the heating module, having a slot, made with a CNC milling machine, to insert the disc. Heater mats with the attached copper plates in the center were attached on either side of the PMMA sheet, thus forming a heating chamber for the disc with an open slit on the side for inserting disc. The thickness of the copper plates and PMMA sheets was chosen to ensure direct contact with the two copper plates upon insertion of the disc into the chamber. Finally, the heating chamber was closed to ensure rapid and uniform heating using a PMMA part cut to fit into the slit opening. The heating module was lined with insulating foam on either side and placed between two 4 mm thick PMMA sheets (150 × 250 mm) which were then closed using M6 bolts on the 4 corners. A thermocouple (Type K; RS PRO Article # 621-2170) was embedded between the copper plate and the heater mat to measure the temperature. Temperature regulation was achieved by using N-channel MOSFET (IRF540PBF, 28A, 100V; RS PRO article # 708-5143) in combination with Arduino UNO Rev 3 MCU Development Board controlling the power supply to the heater mats.

### 4.3. Assembly of the centrifugal and signal acquisition modules

The centrifugal module (140 × 230 × 80 mm) was fabricated using 4 mm thick PMMA sheets which were cut using CNC milling machine and assembled using M3 bolts and nuts. It consisted of two chambers; the bottom chamber served as the housing for the electronic components and the top chamber is where the rotation of the disc and the signal acquisition was done. The control of the whole platform is achieved using an Arduino UNO Rev 3 MCU Development Board. In the center of the top chamber there was a motor (Emax MT-2204; 2300 kv brushless DC motor) used to rotate the disc. A small piece of magnetic tape was attached to the body of the motor and a bipolar hall effect sensor (Honeywell SS411A; RS Pro Article # 181-1463) was used to measure the rotation speed in real time while sending feedback to the Arduino board. The top chamber also included a blue laser diode (Osram Opto PL 450 nm, 80 mW; RS PRo Article # 758-7810) which was used for signal acquisition. A 16×2 pixel display module attached at the front provides information to the user regarding ongoing assay steps. Further technical details and exploded view is shown in **Figure S4**.

### 4.4. Packing, drying and characterization of N-benzyl-N-methylethanolamine (NBNM) agarose beads

The NBNM agarose beads were packed in the discs by first diluting the bead resin stock (Capto Adhere, Cytiva) in DI-water at 10% (v/v). 20 μL of the diluted bead stock were then added to each channel on the disc and subsequently centrifuged at 6000 rpm for 10 s. The excess solution was manually removed from the outlet and the disc was subsequently placed in a vacuum chamber at 15 Pa for 8 min to dehydrate the beads. After dehydration, the disc was centrifuged a second time at 6000 rpm for 10 s to ensure a homogeneous packing. The bp-cutoff of the beads packed inside the discs was characterized by microfluidic capillary electrophoresis in a Bioanalyzer 2100 system with a DNA 1000 kit (Agilent Technologies, USA).

The packing in the PDMS microchannels was performed by first preparing a suspension of beads in a 20% PEG 8000 (w/w) solution by adding 1 μL bead stock to 19 μL PEG solution. The suspension was flowed into the microchannels at 10 μL/min using a NE-1200 syringe pump (New Era Pump Systems, USA). The channels were then rinsed with DI-water flowed at 10 μL/min for 5 min to ensure removal of any residual PEG and salts. To dehydrate the beads, the PDMS device was placed in a vacuum chamber at 15 Pa for 5 min and subsequently stored at room temperature until further usage. The beads inside the PDMS devices were characterized by fluorescence microscopy in a Nikon Ti-Eclipse inverted microscope equipped with a Lumencor SOLA light engine and a FITC filter cube. The acquired fluorescence microscopy images were analyzed using ImageJ software (NIH, USA).

### 4.5. Real-time loop mediated isothermal amplification (RT-LAMP) of SARS-CoV-2 RNA

The RT-LAMP was performed in a Mic qPCR Cycler (Biomolecular systems, Australia) with a total of 10 μL per reaction mixture. The reaction mixture containing 1 μL template RNA was prepared with final concentrations of 1x Isothermal amplification buffer II (New England Biolabs, USA), 6 mM dNTPs (RP65, Blirt, Poland), 6 mM MgSO_4_ (New England Biolabs, USA), 1x primer mixture (six primers described in detail below), 1x SYBR Green I (S7585, Thermo Fisher Scientific), 320 mU Bst 3.0 polymerase (New England Biolabs, USA) and 4 U SuperScript IV reverse transcriptase (Thermo Fisher Scientific). The primer mixture at 10x concentration was prepared with 2 μM *F* and *B*, 16 μM *FIP* and *BIP* and 4 μM *LF* and *LB*. The primers sequences were the following: *F*-5’-CCA CTA GAG GAG CTA CTG TA-3’; *B*- 5’-TGA CAA GCT ACA ACA CGT-3’; *FIP*-5’-AGG TGA GGG TTT TCT ACA TCA CTA TAT TGG AAC AAG CAA ATT CTA TGG-3’; *BIP*- 5’-ATG GGT TGG GAT TAT CCT AAA TGT GTG CGA GCA AGA ACA AGT G-3’; *LF*- 5’-CAG TTT TTA ACA TGT TGT GCC AAC C-3’; *LB*- 5’-TAG AGC CAT GCC TAA CAT GCT-3’. All primers were synthesized by Thermo Fisher Scientific. The RNA fragment (226 bp) matching the sequence of the ORF1ab of SARS-CoV-2 (MT883505.1, 15118-15343) used to characterize and optimize the LAMP reaction was produced by in-vitro transcription and stored at −80 °C in DI-water at 5 ng/μL.

### 4.6. Collection and processing of nasopharyngeal swab samples

We obtained nasopharyngeal samples from Karolinska University Hospital, Huddinge (Stockholm) collected between May 20^th^ and June 1^st^ 2020. Samples were collected either using Sigma-Transwab or Sigma-Virocult kits (MWE, UK) as described in detail elsewhere ^46^. We used pseudo-anonymized surplus material previously collected for clinical diagnostics of SARS-CoV-2. This is in accordance with the Swedish Act concerning the Ethical Review of Research Involving Humans, which allows development and improvement of diagnostic assays using patient samples which were collected to perform the testing in question. Additional ethical approval for RT-LAMP diagnosis was obtained by the appropriate Swedish Authority (Dnr 2020-01945, Etikproevningsnaemnden). All samples were pre-analyzed on the GeneXpert Xpress SARS-CoV-2 system (Cepheid, USA) and Ct values for E and N2 genes were obtained. For the LAMP experiments, 50 μl of each sample was heat-inactivated at 95 °C for 15 minutes and stored at −80 °C until further processing.

### 4.7. Detection of SARS-CoV-2 RNA using the integrated centrifugal platform

The sample containing the RNA template (*in vitro* synthetized RNA fragment or inactivated SARS-CoV-2) was first combined with the LAMP master mix at a ratio of 10% (v/v) and 10 μL were subsequently added to each of the 20 channels in the disc. Both inlet and outlet access holes of all channels were sealed with a sheet of PSA (ARSeal™ 90880) covering the entire disc surface. The sealed disc was subsequently inserted into the heater module and incubated at 65 °C for 25-30 min. The disc was then transferred to the centrifugal platform and subjected to 4 acceleration/deceleration cycles from 0 to 6,000 rpm taking 1 min each. After centrifugation, the laser diode was turned on and each channel was sequentially imaged using a mid-range smartphone camera (OnePlus 6T, China). The acquired RGB images were processed using ImageJ software (NIH, USA) measuring the grayscale profile of the green channel along the interface between the solution and the packed beads (100 pixels long and average of 30 pixels perpendicular to the line). The grayscale intensity on the beads was normalized relative to the solution for each channel and was smoothed with a 10-pixel moving average before further processing.

## Data Availability

The authors confirm that the data supporting the findings of this study are available within the article and its supplementary materials.

## Acknowledgements

This work has been sponsored in part by the European Union’s Horizon 2020 research and innovation programme “New Diagnostics for Infectious Diseases” (ND4ID) under the Marie Skłodowska-Curie grant agreement No. 675412. VP is supported by a SciLifeLab/KAW national COVID-19 research program project grant (KAW 2020.0182), Yin is supported by Ganzhou COVID-19 Emergency Research Project, Key Special Project of “Technology Boosts Economy 2020” of Ministry of Science and Technology (SQ2020YFF0411358) for COVID-19 related work. The authors acknowledge Alisa Alekseenko and Yerma Pareja-Sanchez for technical assistance with the RT-LAMP.

## Supporting Information

**Figure S1.**
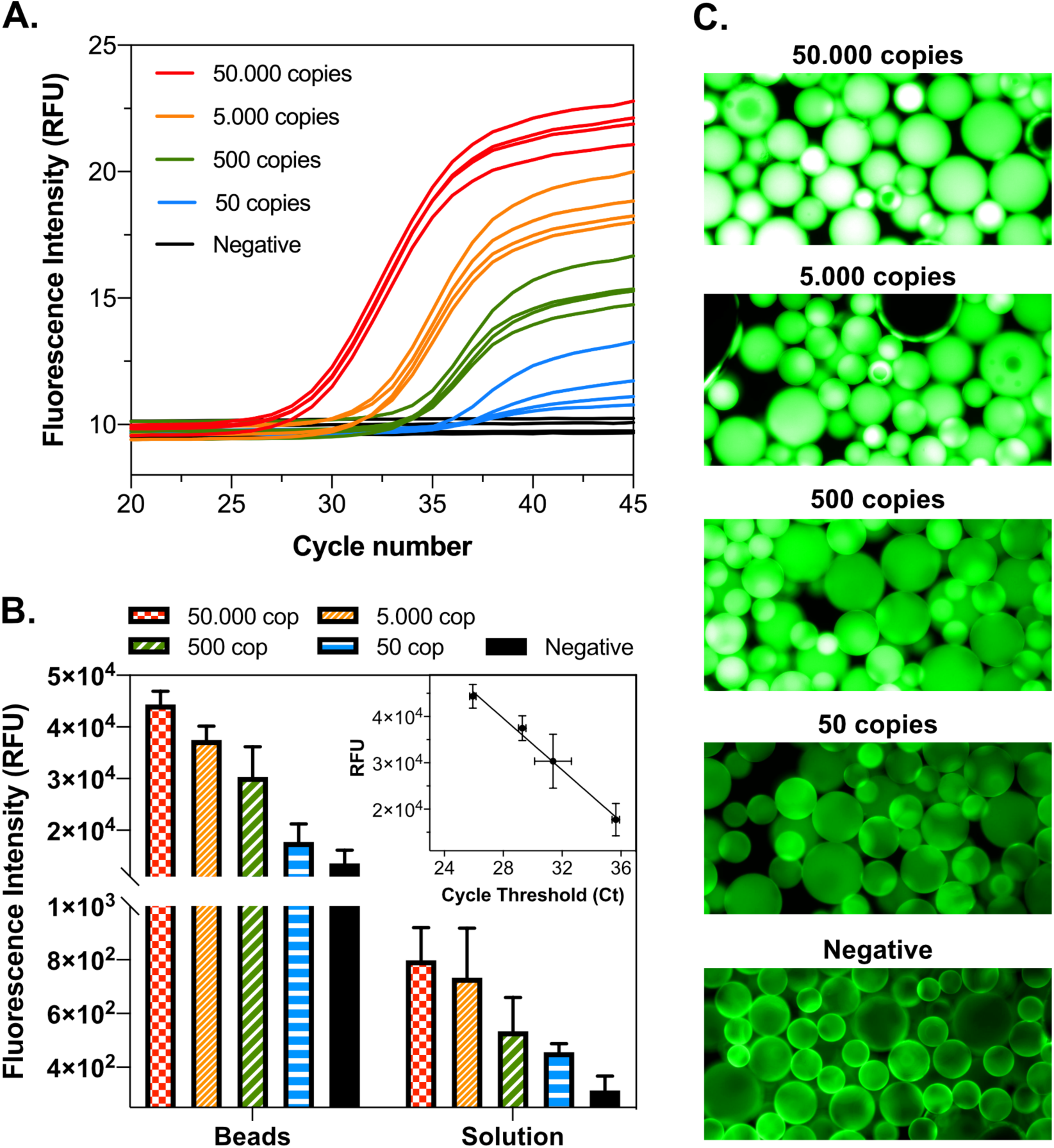
Proof-of-concept of PCR amplicon enrichment on N-benzyl-N-methylethanolamine beads. Synthetic SARS-CoV-2 full genomic RNA (MN908947.3, Twist Bioscience) was spiked at a total of 50 to 50.000 copies per 25 μL reaction mix containing 1x TaqMan FAST Virus 1-step MM (Thermo Fisher Scientific), 0.6 μM forward primer (5’-GTG ARA TGG TCA TGT GTG GCG G-3’), 0.8 μM reverse primer (5’-CAR ATG TTA AAS ACA CTA TTA GCA TA-3’) and 0.2 uM of molecular beacon probe 1 (5’-FAM-CAG GTG GAA CCT CAT CAG GAG ATG C-BHQ_1-3’) and 2 (5’-FAM-CCA GGT GGW ACR TCA TCM GGT GAT GC-BHQ_1-3’). The mixture was first incubated for 5 min at 50 °C for reverse transcription, denatured for 20 s at 95 °C and subjected to a series of 45 cycles of 3 s at 95 °C and 30 s at 60 °C. The amplification generates a 100 bp long amplicon. A-RT-PCR with increasing RNA copy numbers in solution, measured using a micPCR magnetic induction cycler (BMS, Australia). Each viral copy titer was measured in quadruplicate. B-Fluorescence intensity measured on Capto Adhere beads, or solution upstream of the beads after flowing 10 μL of the pre-amplified mixture (after the 45 amplification cycles in A) through the bead-packed microchannel. Both fluorescence intensity on the beads and in solution were measured using fluorescence microscopy and ImageJ software (NIH, USA) by measuring the grey scale intensity in both regions (16-bit images). The beads provide an increase of ∼100-fold in fluorescence signal intensity. Error bars correspond to the standard deviation of 4 independent PCR amplification and bead-capture experiments. The inset plot shows the correlation between cycle threshold values of the tested viral loads and fluorescence intensity measured on the beads. C-Microscopy images of the beads with increasing copy numbers of SARS-CoV-2 genomic RNA (initial copy numbers before PCR).

**Figure S2.**
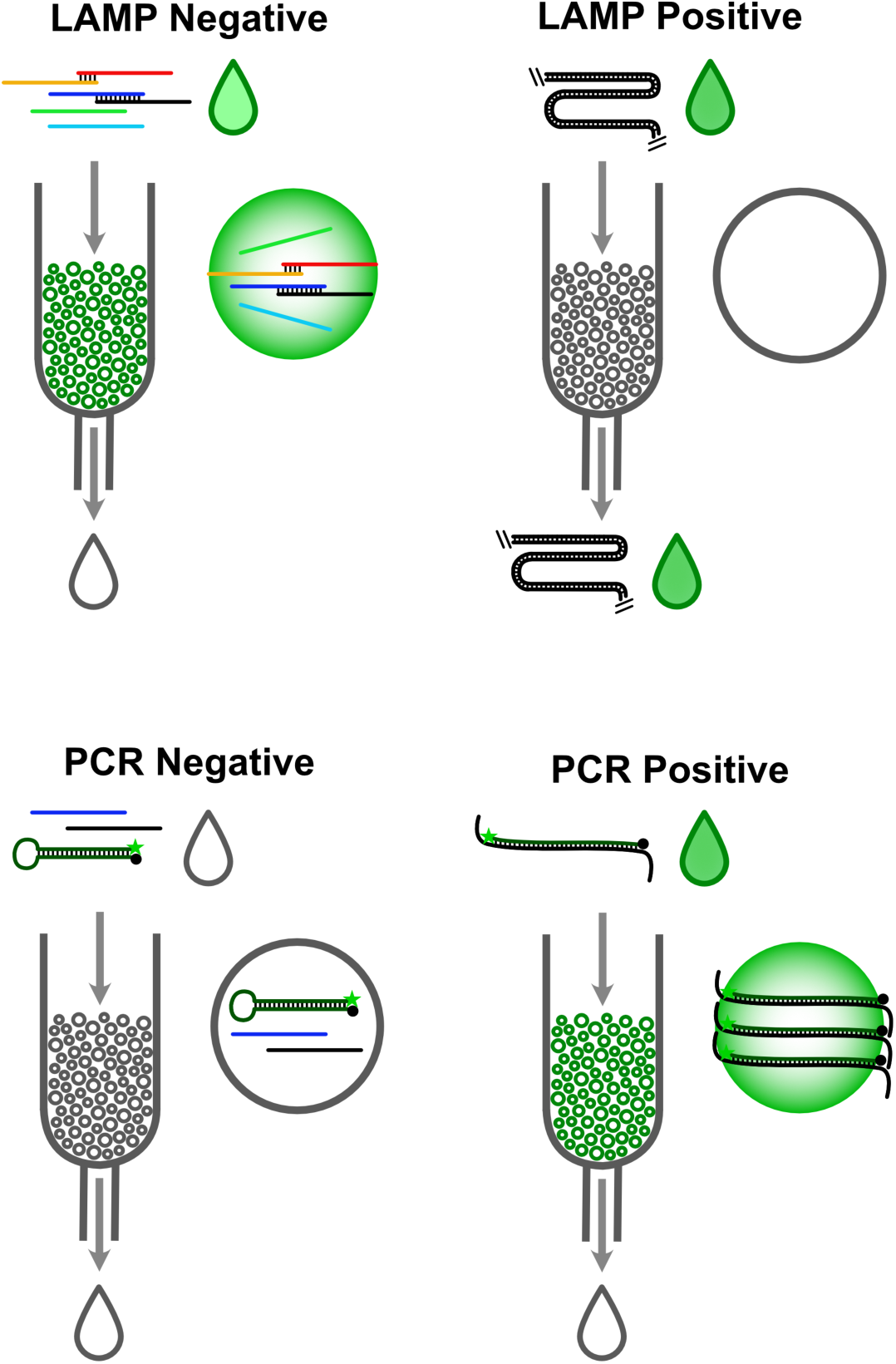
Schematics of the bead capture selectivity in the presence of positive or negative samples amplified with LAMP or PCR. In the case of LAMP, the fluorescence is generated by a dsDNA fluorescent intercalator (e.g. SYBR green I). For PCR, the fluorescence is generated using a molecular beacon (e.g. FAM-BHQ-1 pair). In the tested setup, the PCR amplicon has a length of 100 bp and is able to effectively penetrate the pores of the agarose beads. Green droplets indicate strongly fluorescent solutions, while white droplets indicate non-fluorescent or weakly fluorescent solutions.

**Figure S3.**
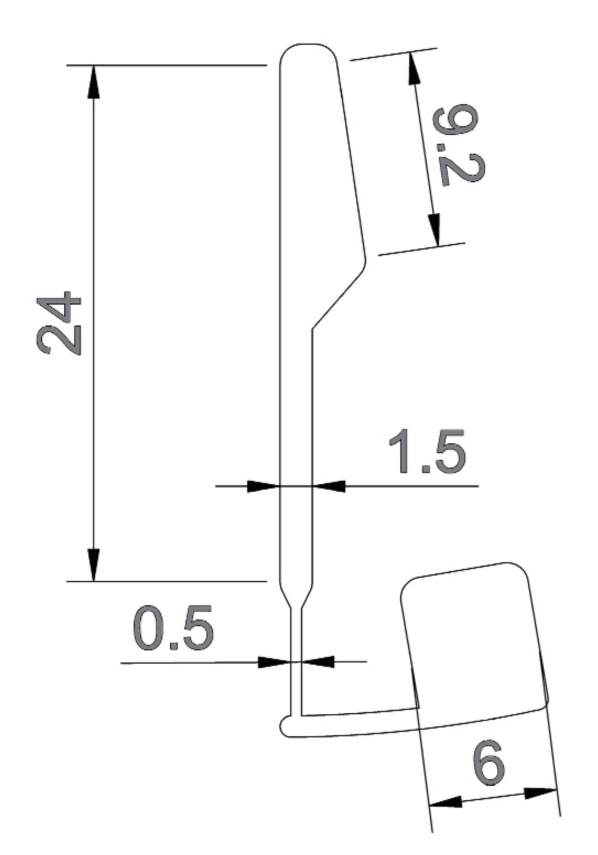
Detailed dimensions (parallel to the rotation axis) of the microchannels drilled on the discs. All values are in mm units. The depth of the channel is 400 μm except at the interface between the 0.5 mm and 6 mm wide regions, where the depth is 50 μm.

**Figure S4.**
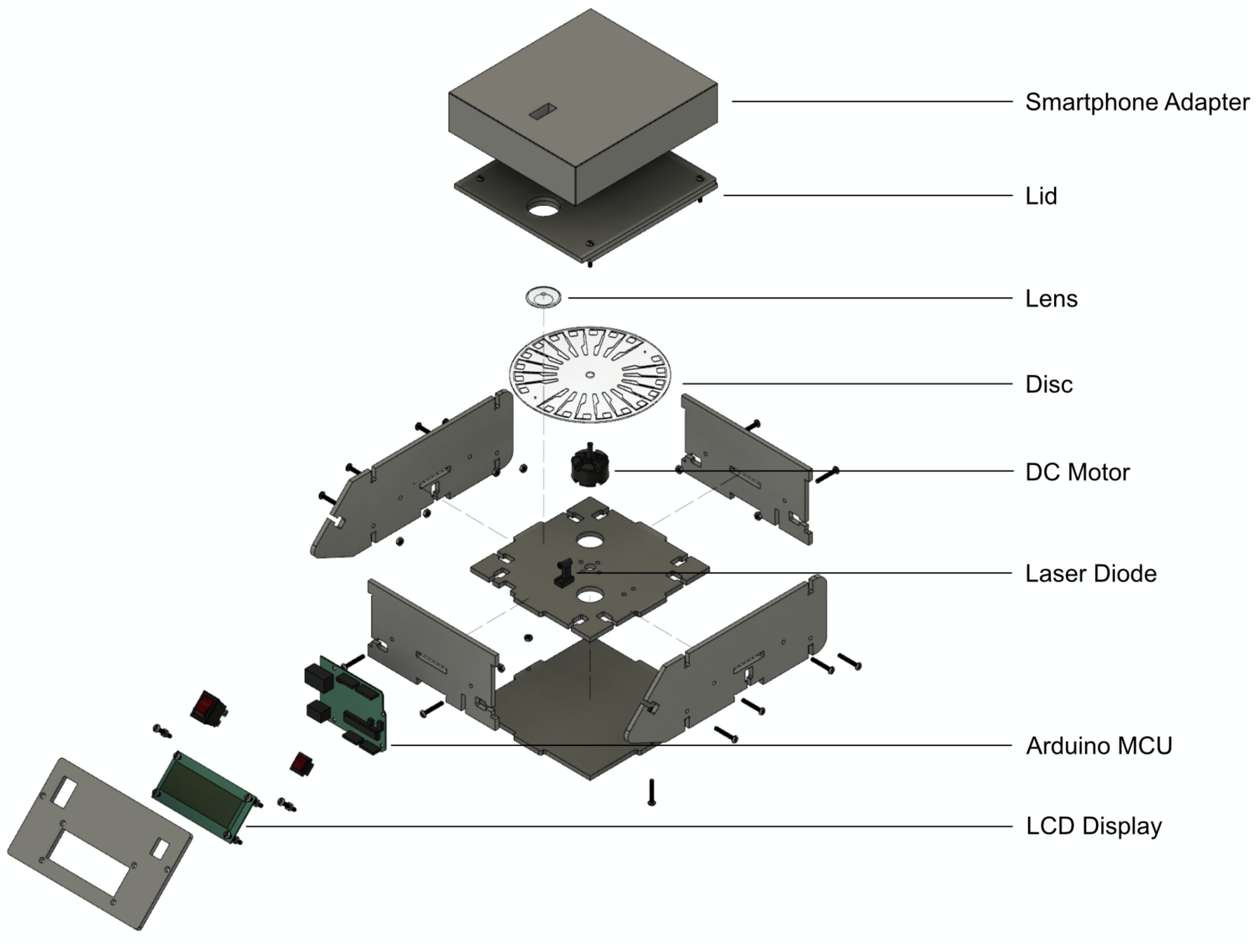
Exploded view of the centrifugal platform. The smartphone adapter design depends on the specific position of the camera being used and can be adapted to ensure an optimal focal distance when measuring the fluorescence signal on the microchannels. A Kapton™ 100 μm thick polyimide film is attached between the lens and the disc to block the excitation light from the laser diode.

**Figure S5.**
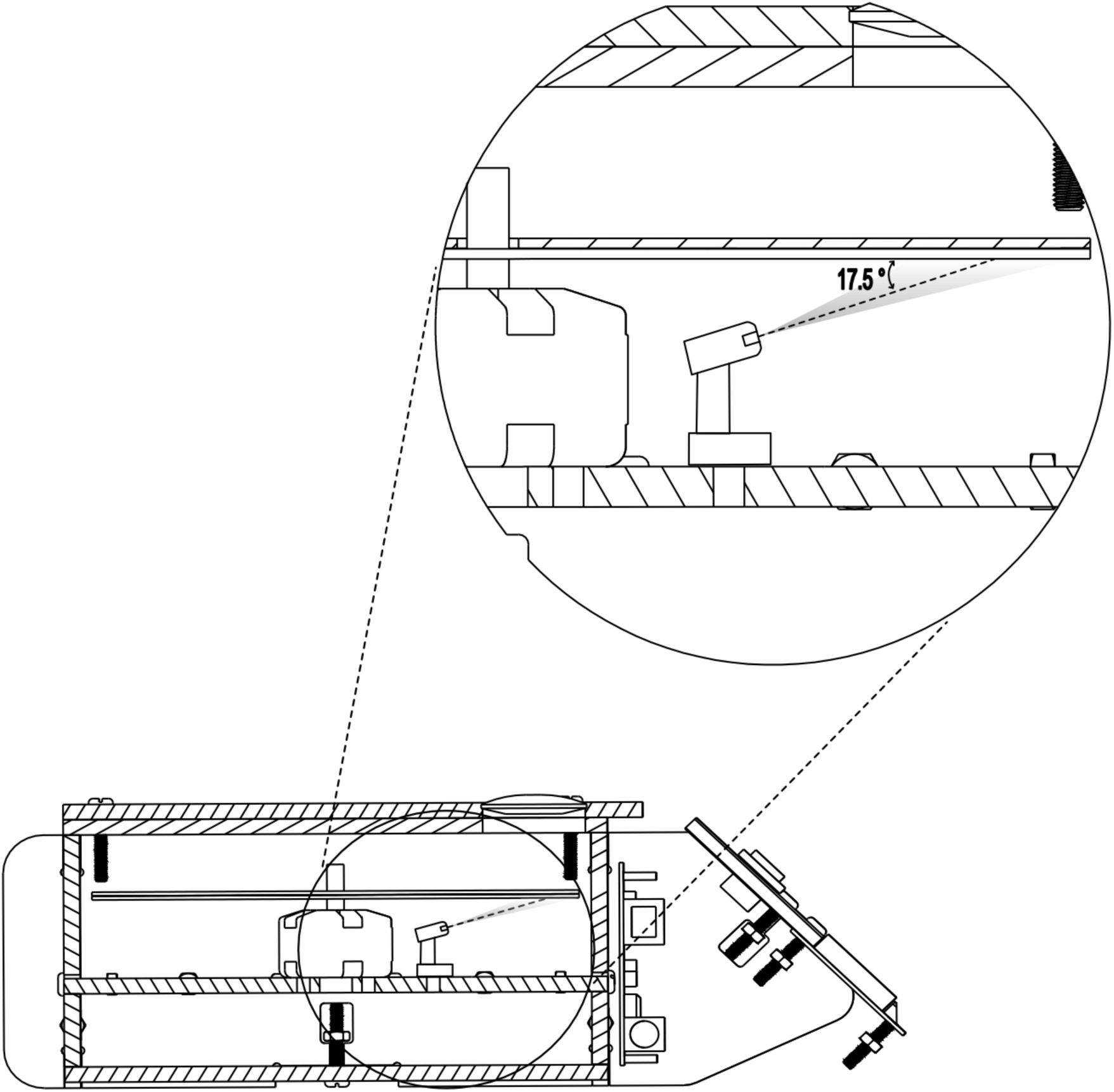
Position and incidence angle of the laser light relative to the disc. The light from the laser has a perpendicular beam divergence of 25° (FWHM) and a parallel beam divergence of 11°. The higher perpendicular divergence ensures illumination of the entire solution and bead region along the microchannels.

## References

(1) Racsa, L. D., Kraft, C. S.; Olinger, G. G.; Hensley, L. E. Viral Hemorrhagic Fever Diagnostics, Clin Infect Dis 2016, 62, 214–219.

(2) Shorten, R. J.; Brown, C. S.; Jacobs, M.; Rattenbury, S.; Simpson, A. J.; Mepham, S. Diagnostics in Ebola Virus Disease in Resource-Rich and Resource-Limited Settings, PLoS Negl Trop Dis 2016, 10, e0004948.

(3) Weidemaier, K.; Carrino, J.; Curry, A.; Connor, J. H.; Liebmann-Vinson, A. Advancing rapid point-of-care viral diagnostics to a clinical setting, Future Virology 2015, 10, 313–328.

(4) MacKay, M. J.; Hooker, A. C.; Afshinnekoo, E.; Salit, M.; Kelly, J.; Feldstein, J. V.; Haft, N.; Schenkel, D.; Nambi, S.; Cai, Y.; Zhang, F.; Church, G.; Dai, J.; Wang, C. L.; Levy, S.; Huber, J.; Ji, H. P.; Kriegel, A.; Wyllie, A. L.; Mason, C. E. The COVID-19 XPRIZE and the need for scalable, fast, and widespread testing, Nat. Biotechnol. 2020.

(5) Lu, R.; Zhao, X.; Li, J.; Niu, P.; Yang, B.; Wu, H.; Wang, W.; Song, H.; Huang, B.; Zhu, N.; Bi, Y.; Ma, X.; Zhan, F.; Wang, L.; Hu, T.; Zhou, H.; Hu, Z.; Zhou, W.; Zhao, L.; Chen, J.; et al. Genomic characterisation and epidemiology of 2019 novel coronavirus: implications for virus origins and receptor binding, The Lancet 2020, 395, 565–574.

(6) To, K. K.-W.; Tsang, O. T.-Y.; Leung, W.-S.; Tam, A. R.; Wu, T.-C.; Lung, D. C.; Yip, C. C.-Y.; Cai, J.-P.; Chan, J. M.-C.; Chik, T. S.-H.; Lau, D. P.-L.; Choi, C. Y.-C.; Chen, L.-L.; Chan, W.-M.; Chan, K.-H.; Ip, J. D.; Ng, A. C.-K.; Poon, R. W.-S.; Luo, C.-T.; Cheng, V. C.-C.; et al. Temporal profiles of viral load in posterior oropharyngeal saliva samples and serum antibody responses during infection by SARS-CoV-2: an observational cohort study, The Lancet Infectious Diseases 2020, 20, 565–574.

(7) Pan, Y.; Zhang, D.; Yang, P.; Poon, L. L. M.; Wang, Q. Viral load of SARS-CoV-2 in clinical samples, The Lancet Infectious Diseases 2020, 20, 411–412.

(8) Li, R.; Pei, S.; Chen, B.; Song, Y.; Zhang, T.; Yang, W.; Shaman, J. Substantial undocumented infection facilitates the rapid dissemination of novel coronavirus (SARS-CoV-2), Science 2020, 368, 489–493.

(9) Walsh, K. A.; Jordan, K.; Clyne, B.; Rohde, D.; Drummond, L.; Byrne, P.; Ahern, S.; Carty, P. G.; O’Brien, K. K.; O’Murchu, E.; O’Neill, M.; Smith, S. M.; Ryan, M.; Harrington, P. SARS-CoV-2 detection, viral load and infectivity over the course of an infection, J Infect 2020, 81, 357–371.

(10) Waggoner, J. J.; Stittleburg, V.; Pond, R.; Saklawi, Y.; Sahoo, M. K.; Babiker, A.; Hussaini, L.; Kraft, C. S.; Pinsky, B. A.; Anderson, E. J.; Rouphael, N. Triplex Real-Time RT-PCR for Severe Acute Respiratory Syndrome Coronavirus 2, Emerg Infect Dis 2020, 26, 1633–1635.

(11) Wang, X.; Yao, H.; Xu, X.; Zhang, P.; Zhang, M.; Shao, J.; Xiao, Y.; Wang, H. Limits of Detection of 6 Approved RT–PCR Kits for the Novel SARS-Coronavirus-2 (SARS-CoV-2), Clin. Chem. 2020, 66, 977–979.

(12) Corman, V. M.; Landt, O.; Kaiser, M.; Molenkamp, R.; Meijer, A.; Chu, D. K.; Bleicker, T.; Brunink, S.; Schneider, J.; Schmidt, M. L.; Mulders, D. G.; Haagmans, B. L.; van der Veer, B.; van den Brink, S.; Wijsman, L.; Goderski, G.; Romette, J. L.; Ellis,J., Zambon, M.; Peiris, M.; et al. Detection of 2019 novel coronavirus (2019-nCoV) by real-time RT-PCR, Euro Surveill 2020, 25.

(13) Yu, F.; Yan, L.; Wang, N.; Yang, S.; Wang, L.; Tang, Y.; Gao, G.; Wang, S.; Ma, C.; Xie, R.; Wang, F.; Tan, C.; Zhu, L.; Guo, Y.; Zhang, F. Quantitative Detection and Viral Load Analysis of SARS-CoV-2 in Infected Patients, Clin Infect Dis 2020, 71, 793–798.

(14) Wolfel, R.; Corman, V. M.; Guggemos, W.; Seilmaier, M.; Zange, S.; Muller, M. A.; Niemeyer, D.; Jones, T. C.; Vollmar, P.; Rothe, C.; Hoelscher, M.; Bleicker, T.; Brunink, S.; Schneider, J.; Ehmann, R.; Zwirglmaier, K.; Drosten, C.; Wendtner, C. Virological assessment of hospitalized patients with COVID-2019, Nature 2020, 581, 465–469.

(15) Kampe, J. J. A. v.; Vijver, D. A. M. C. v. d.; Fraaij, P. L. A.; Haagmans, B. L.; Lamers, M. M.; Okba, N.; Akker, J. P. C. v. d.; Endeman, H.; Gommers, D. A. M. P. J.; Cornelissen, J. J.; Hoek, R. A. S.; der, M. M. v.; Eerden Hesselink, D.A., Metselaar, H. J.; Verbon, A.; Steenwinkel, J. E. M. d.; Aron, G. I.; Gorp, E. C. M. v.; Boheemen, S. v.; et al. Shedding of infectious virus in hospitalized patients with coronavirus disease-2019 (COVID-19) duration and key determinants, MedRxiv 2020.

(16) Mina, M. J.; Parker, R.; Larremore, D. B. Rethinking Covid-19 Test Sensitivity — A Strategy for Containment, New England Journal of Medicine 2020.

(17) Dinnes, J.; Deeks, J. J.; Adriano, A.; Berhane, S.; Davenport, C.; Dittrich, S.; Emperador, D.; Takwoingi, Y.; Cunningham, J.; Beese, S.; Dretzke, J.; Ferrante di Ruffano, L., Harris, I. M.; Price, M. J.; Taylor-Phillips, S.; Hooft, L.; Leeflang, M. M.; Spijker, R.; Van den Bruel, A.; Cochrane, C.-D. T. A. G. Rapid, point-of-care antigen and molecular-based tests for diagnosis of SARS-CoV-2 infection, Cochrane Database Syst Rev 2020, 8, CD013705.

(18) Kalk, A.; Schultz, A. SARS-CoV-2 epidemic in African countries—are we losing perspective?, The Lancet Infectious Diseases 2020.

(19) Zhang, H.; Xu, Y.; Fohlerova, Z.; Chang, H.; Iliescu, C.; Neuzil, P. LAMP-on-a-chip: Revising microfluidic platforms for loop-mediated DNA amplification, Trends Analyt Chem 2019, 113, 44–53.

(20) Oh, S. J.; Park, B. H.; Jung, J. H.; Choi, G.; Lee, D. C.; Kim, D. H.; Seo, T. S. Centrifugal loop-mediated isothermal amplification microdevice for rapid, multiplex and colorimetric foodborne pathogen detection, Biosens. Bioelectron. 2016, 75, 293–300.

(21) Park, B. H.; Oh, S. J.; Jung, J. H.; Choi, G.; Seo, J. H.; Kim, D. H.; Lee, E. Y.; Seo, T. S. An integrated rotary microfluidic system with DNA extraction, loop-mediated isothermal amplification, and lateral flow strip based detection for point-of-care pathogen diagnostics, Biosens. Bioelectron. 2017, 91, 334–340.

(22) Yan, H.; Zhu, Y.; Zhang, Y.; Wang, L.; Chen, J.; Lu, Y.; Xu, Y.; Xing, W. Multiplex detection of bacteria on an integrated centrifugal disk using bead-beating lysis and loop-mediated amplification, Sci Rep 2017, 7, 1460.

(23) Sayad, A. A.; Ibrahim, F.; Uddin, S. M.; Pei, K. X.; Mohktar, M. S.; Madou, M.; Thong, K. L. A microfluidic lab-on-a-disc integrated loop mediated isothermal amplification for foodborne pathogen detection, Sens. Actuators, B 2016, 227, 600–609.

(24) Liu, D.; Zhu, Y.; Li, N.; Lu, Y.; Cheng, J.; Xu, Y. A portable microfluidic analyzer for integrated bacterial detection using visible loop-mediated amplification, Sens. Actuators, B 2020, 310.

(25) Yamanaka, E. S.; Tortajada-Genaro, L. A.; Pastor, N.; Maquieira, A. Polymorphism genotyping based on loop-mediated isothermal amplification and smartphone detection, Biosens. Bioelectron. 2018, 109, 177–183.

(26) Wang, H.; Ma, Z.; Qin, J.; Shen, Z.; Liu, Q.; Chen, X.; Wang, H.; An, Z.; Liu, W.; Li, M. A versatile loop-mediated isothermal amplification microchip platform for Streptococcus pneumoniae and Mycoplasma pneumoniae testing at the point of care, Biosens. Bioelectron. 2019, 126, 373–380.

(27) Wan, L.; Gao, J.; Chen, T.; Dong, C.; Li, H.; Wen, Y. Z.; Lun, Z. R.; Jia, Y.; Mak, P. I.; Martins, R. P. LampPort: a handheld digital microfluidic device for loop-mediated isothermal amplification (LAMP), Biomed. Microdevices 2019, 21, 9.

(28) Jackson, K. R.; Layne, T.; Dent, D. A.; Tsuei, A.; Li, J.; Haverstick, D. M.; Landers, J. P. A novel loop-mediated isothermal amplification method for identification of four body fluids with smartphone detection, Forensic Sci Int Genet 2020, 45, 102195.

(29) Hui, J.; Gu, Y.; Zhu, Y.; Chen, Y.; Guo, S. J.; Tao, S. C.; Zhang, Y.; Liu, P. Multiplex sample-to-answer detection of bacteria using a pipette-actuated capillary array comb with integrated DNA extraction, isothermal amplification, and smartphone detection, Lab Chip 2018, 18, 2854–2864.

(30) Wang, S.; Liu, N.; Zheng, L.; Cai, G.; Lin, J. A lab-on-chip device for the sample-in-result-out detection of viable Salmonella using loop-mediated isothermal amplification and real-time turbidity monitoring, Lab Chip 2020, 20, 2296–2305.

(31) Kaarj, K.; Akarapipad, P.; Yoon, J. Y. Simpler, Faster, and Sensitive Zika Virus Assay Using Smartphone Detection of Loop-mediated Isothermal Amplification on Paper Microfluidic Chips, Sci Rep 2018, 8, 12438.

(32) Liao, S.-C.; Peng, J.; Mauk, M. G.; Awasthi, S.; Song, J.; Friedman, H.; Bau, H. H.; Liu, C. Smart cup: A minimally-instrumented, smartphone-based point-of-care molecular diagnostic device, Sens. Actuators, B 2016, 229, 232–238.

(33) Yu, L.; Wu, S.; Hao, X.; Dong, X.; Mao, L.; Pelechano, V.; Chen, W.-H.; Yin, X. Rapid Detection of COVID-19 Coronavirus Using a Reverse Transcriptional Loop-Mediated Isothermal Amplification (RT-LAMP) Diagnostic Platform, Clin. Chem. 2020, 66, 975–977.

(34) Yan, C.; Cui, J.; Huang, L.; Du, B.; Chen, L.; Xue, G.; Li, S.; Zhang, W.; Zhao, L.; Sun, Y.; Yao, H.; Li, N.; Zhao, H.; Feng, Y.; Liu, S.; Zhang, Q.; Liu, D.; Yuan, J. Rapid and visual detection of 2019 novel coronavirus (SARS-CoV-2) by a reverse transcription loop-mediated isothermal amplification assay, Clin Microbiol Infect 2020, 26, 773–779.

(35) Baek, Y. H.; Um, J.; Antigua, K. J. C.; Park, J. H.; Kim, Y.; Oh, S.; Kim, Y. I.; Choi, W. S.; Kim, S. G.; Jeong, J. H.; Chin, B. S.; Nicolas, H. D. G.; Ahn, J. Y.; Shin, K. S.; Choi, Y. K.; Park, J. S.; Song, M. S. Development of a reverse transcription-loop-mediated isothermal amplification as a rapid early-detection method for novel SARS-CoV-2, Emerg Microbes Infect 2020, 9, 998–1007.

(36) Park, G. S.; Ku, K.; Baek, S. H.; Kim, S. J.; Kim, S. I.; Kim, B. T.; Maeng, J. S. Development of Reverse Transcription Loop-Mediated Isothermal Amplification Assays Targeting Severe Acute Respiratory Syndrome Coronavirus 2 (SARS-CoV-2), J Mol Diagn 2020, 22, 729–735.

(37) Huang, W. E.; Lim, B.; Hsu, C. C.; Xiong, D.; Wu, W.; Yu, Y.; Jia, H.; Wang, Y.; Zeng, Y.; Ji, M.; Chang, H.; Zhang, X.; Wang, H.; Cui, Z. RT-LAMP for rapid diagnosis of coronavirus SARS-CoV-2, Microb Biotechnol 2020, 13, 950–961.

(38) Lamb, L. E.; Bartolone, S. N.; Ward, E.; Chancellor, M. B. Rapid detection of novel coronavirus/Severe Acute Respiratory Syndrome Coronavirus 2 (SARS-CoV-2) by reverse transcription-loop-mediated isothermal amplification, PloS one 2020, 15, e0234682.

(39) Thi, V. L. D.; Herbst, K.; Boerner, K.; Meurer, M.; Kremer, L. P.; Kirrmaier, D.; Freistaedter, A.; Papagiannidis, D.; Galmozzi, C.; Stanifer, M. L.; Boulant, S.; Klein, S.; Chlanda, P.; Khalid, D.; Miranda, I. B.; Schnitzler, P.; Kräusslich, H.-G.; Knop, M.,Anders, S. A colorimetric RT-LAMP assay and LAMP-sequencing for detecting SARS-CoV-2 RNA in clinical samples, Science Translational Medicine 2020, 12, eabc7075.

(40) Novo, P.; Chu, V.; Conde, J. P. Integrated fluorescence detection of labeled biomolecules using a prism-like PDMS microfluidic chip and lateral light excitation, Lab Chip 2014.

(41) Meagher, R. J.; Priye, A.; Light, Y. K.; Huang, C.; Wang, E. Impact of primer dimers and self-amplifying hairpins on reverse transcription loop-mediated isothermal amplification detection of viral RNA, Analyst 2018, 143, 1924–1933.

(42) Pinto, I. F.; Aires-Barros, M. R.; Azevedo, A. M. Multimodal chromatography: debottlenecking the downstream processing of monoclonal antibodies, Pharmaceutical Bioprocessing 2015, 3, 263–279.

(43) Matos, T.; Queiroz, J. A.; Bülow, L. Plasmid DNA purification using a multimodal chromatography resin, Journal of Molecular Recognition 2014, 27, 184–189.

(44) Carturan, S.; Quaranta, A.; Bonafini, M.; Vomiero, A.; Maggioni, G.; Mattei, G.; de Julián Fernández, C.; Bersani, M.; Mazzoldi, P.; Della Mea, G. Formation of silver nanoclusters in transparent polyimides by Ag-K ion-exchange process, The European Physical Journal D 2007, 42, 243–251.

(45) Wolters, F.; van de Bovenkamp, J.; van den Bosch, B.; van den Brink, S.; Broeders, M.; Chung, N. H.; Favie, B.; Goderski, G.; Kuijpers, J.; Overdevest, I.; Rahamat-Langedoen, J.; Wijsman, L.; Melchers, W. J.; Meijer, A. Multi-center evaluation of cepheid xpert(R) xpress SARS-CoV-2 point-of-care test during the SARS-CoV-2 pandemic, J Clin Virol 2020, 128, 104426.

(46) Alekseenko, A.; Barrett, D.; Pareja-Sanchez, Y.; Howard, R. J.; Strandback, E.; Ampah-Korsah, H.; Rovšnik, U.; Zuniga-Veliz, S.; Klenov, A.; Malloo, J.; Ye, S.; Liu, X.; Reinius, B.; Elsässer, S.; Nyman, T.; Sandh, G.; Yin, X.; Pelechano, V. Detection of SARS-CoV-2 using non-commercial RT-LAMP reagents and raw samples, medRxiv 2020, 2020.08.22.20179507.

(47) Pinto, I. F.; Caneira, C. R. F.; Soares, R. R. G.; Madaboosi, N.; Aires-Barros, M. R.; Conde, J. P.; Azevedo, A. M., Chu, V. The application of microbeads to microfluidic systems for enhanced detection and purification of biomolecules, Methods 2017, 116, 112–124.

